# Automated assessment of neonatal internal capsule maturation on T2-weighted MRI across 7T and 3T

**DOI:** 10.64898/2026.06.02.26354741

**Authors:** Chiara Casella, Alena Uus, Luke Dedominicis, Jucha Willers Moore, Benjamin Clayden, Emil Galanides, Philippa Bridgen, Pierluigi Di Cio, Ines Tomazinho, Cidalia Da Costa, Dario Gallo, Sophie Arulkumaran, Maria Deprez, Serena J. Counsell, A. David Edwards, Joseph V. Hajnal, Jonathan O’Muircheartaigh, Mary A. Rutherford, Shaihan Malik, Tomoki Arichi

**Author notes:** Email address:* (Chiara Casella). Joint first authorship C.C. and A.U.

## Abstract

**Motivation:** Quantitative assessment of neonatal internal capsule (IC) maturation remains largely reliant on qualitative visual evaluation, limiting objectivity and scalability.

**Approach:** We developed a fully automated 3D deep learning framework for anatomically detailed segmentation of IC subregions and PLIC myelin-related signal from structural T2-weighted MRI, trained on both high-resolution 7T and conventional 3T neonatal datasets. Volumetric and intensity-based metrics were derived, and developmental trajectories were modelled using postmenstrual age (PMA) and postnatal age (PNA), with normative modelling used to quantify individual deviations.

**Results:** The pipeline achieved high segmentation accuracy across field strengths (Dice > 0.95, relative volume difference < 5%). IC metrics showed robust age-related changes, with volumetric measures increasing and intensity-based measures decreasing with PMA. PNA effects indicated prematurity-related modulation at equivalent maturational age. These patterns generalized to 3T, where normative modelling revealed significant deviations in preterm infants, particularly for myelin-related intensity measures.

**Conclusion:** Structural T2-weighted MRI, combined with anatomically informed segmentation, enables quantitative and biologically meaningful assessment of neonatal IC maturation. This provides a scalable framework for studying early white matter development and supports potential clinical translation.

## 1. Introduction

The internal capsule (IC) is a key subcortical white matter structure located deep and medially within each cerebral hemisphere. It contains densely myelinated ascending and descending fibre tracts that course between the basal ganglia, thalamus, cerebral cortex, brainstem, and spinal cord, forming a critical bidirectional pathway for motor and sensory information (Emos et al., 2023).

The IC is one of the earliest tracts to myelinate during late gestation and early neonatal life (Hasegawa et al., 1992), making it particularly sensitive to maturational changes and vulnerable to perinatal injury, including in infants born preterm (Rutherford et al., 2010). Alterations in T2-weighted (T2w) or T1-weighted (T1w) signal intensity in the posterior limb of the internal capsule (PLIC), assessed at term-equivalent age or by 44 weeks postmenstrual age (PMA), provide a robust marker of both normal maturation and pathology (Counsell et al., 2002b; Rutherford et al., 2010). These signal changes reflect underlying microstructural development, including myelination, and are highly predictive of later motor outcome (Rutherford et al., 1998). Overall, this highlights the IC’s central role as a sensitive locus for detecting injury with long-term functional relevance.

Conventional radiological assessment of IC maturation on T2w and T1w structural MRI typically relies on qualitative visual grading of myelination within its posterior limb (PLIC), which contains fibres of the posterior thalamic radiation, corticospinal, corticorubral, and corticopontine tracts (ARA and ISLAM, 2010). Because the T2w signal is influenced by multiple microstructural factors, including water content and tissue organization, this assessment provides an indirect, albeit clinically established, indication of myelination-related change during early development. Accurate evaluation of PLIC myelination therefore requires substantial specialist expertise, particularly in early developmental stages (before 37 weeks) or acute stages of injury where signal changes might be subtle. In addition, visual assessment in medical imaging is labour-intensive and time-consuming, and is prone to error with substantial inter-observer variability, especially for small structures and suboptimal image contrast or quality.

Recent work has shown promising results for automated quantification of myelination in structural neonatal MRI. In (Wang et al., 2019), thresholding and expectation–maximization were used to segment hypointense ROIs on T2w images, including ventrolateral and subthalamic nuclei as well as myelination within the PLIC. In (Gruber et al., 2022), a multi-view 2D deep learning approach was used to segment the PLIC myelination region on T1-weighted images. However, because these methods focus on signal-defined myelinated regions rather than the full IC, they do not enable regional IC parcellation or anatomical quantification of IC volumes alongside myelination-related signal.

Diffusion MRI–derived metrics such as fractional anisotropy (FA) have been widely used to characterize microstructural properties of the developing brain, including myelination-related changes, and to predict neurodevelopmental outcomes (Dubois et al., 2014; Batalle et al., 2017). However, these approaches typically require dedicated diffusion imaging protocols and complex processing pipelines, which can limit their routine clinical use, particularly in neonatal populations. This creates a need for methods that can derive biologically meaningful markers of maturation from standard structural imaging alone. Ultrahigh-field 7T MRI is particularly promising in this regard, as it provides superior spatial resolution and tissue contrast for visualizing neonatal white matter structures, revealing sharp IC boundaries and subregional features that are indistinct or invisible at conventional 1.5T or 3T field strengths ( Bridgen et al., 2023). However, no established framework currently uses 7T structural MRI to quantitatively characterize IC maturation.

In this work, we developed a deep learning-based seg-mentation pipeline for the internal capsule (IC) using high-resolution 7T MRI, enabling simultaneous quantification of regional volumes and myelination-related intensity metrics from structural T2w imaging alone. Our central aim was to determine whether structural-only MRI can provide anatomically meaningful markers of IC maturation without requiring manual labelling or additional quantitative imaging.

We applied this framework to 7T data to characterize developmental trajectories of IC maturation, and subsequently extended it to conventional 3T neonatal MRI to evaluate its robustness and generalizability in a clinically accessible setting. Within the 3T cohort, we further performed manual PLIC biometry using a template-guided approach to test whether segmentation-derived metrics capture anatomical features routinely assessed in radiological practice. Finally, we applied normative modelling to move from group-level trajectories to individual-level inference, estimating typical development in term-born infants and quantifying subject-specific deviations in preterm infants, thereby testing whether preterm birth is associated with increased deviation from the normative distribution.

## 2. Methods

### 2.1. Cohorts, datasets and preprocessing

Eighty-four neonates (94 scans, including 10 infants with repeat scans; median PMA at scan: 39.00 weeks, range: 33.57–46.71; median GA at birth: 35.14 weeks, range: 27.71–41.57; 52 males) (Fig.1) were imaged during natural sleep with a 7T system (MAGNETOM Terra, Siemens Healthineers, Germany), a 1TX-32RX Nova Medical head coil (Wilmington, MA, USA), and under local safety conditions (REC: RE/LO/1384) (Bridgen et al., 2023; Malik et al., 2021). 2D-TSE sequences (TE = 156 ms, acquired resolution = 0.6 mm, slice thickness = 1.2 mm) were acquired in at least two orthogonal planes, and a 3D volume with 0.45 mm isotropic resolution was created using slice-to-volume reconstruction (Kuklisova-Murgasova et al., 2012). The cohort comprised a heterogeneous sample recruited for research purposes, including healthy controls, preterm neonates, and infants with a range of clinical conditions. Clinical details are reported in Tab. 1.

**Figure 1:**
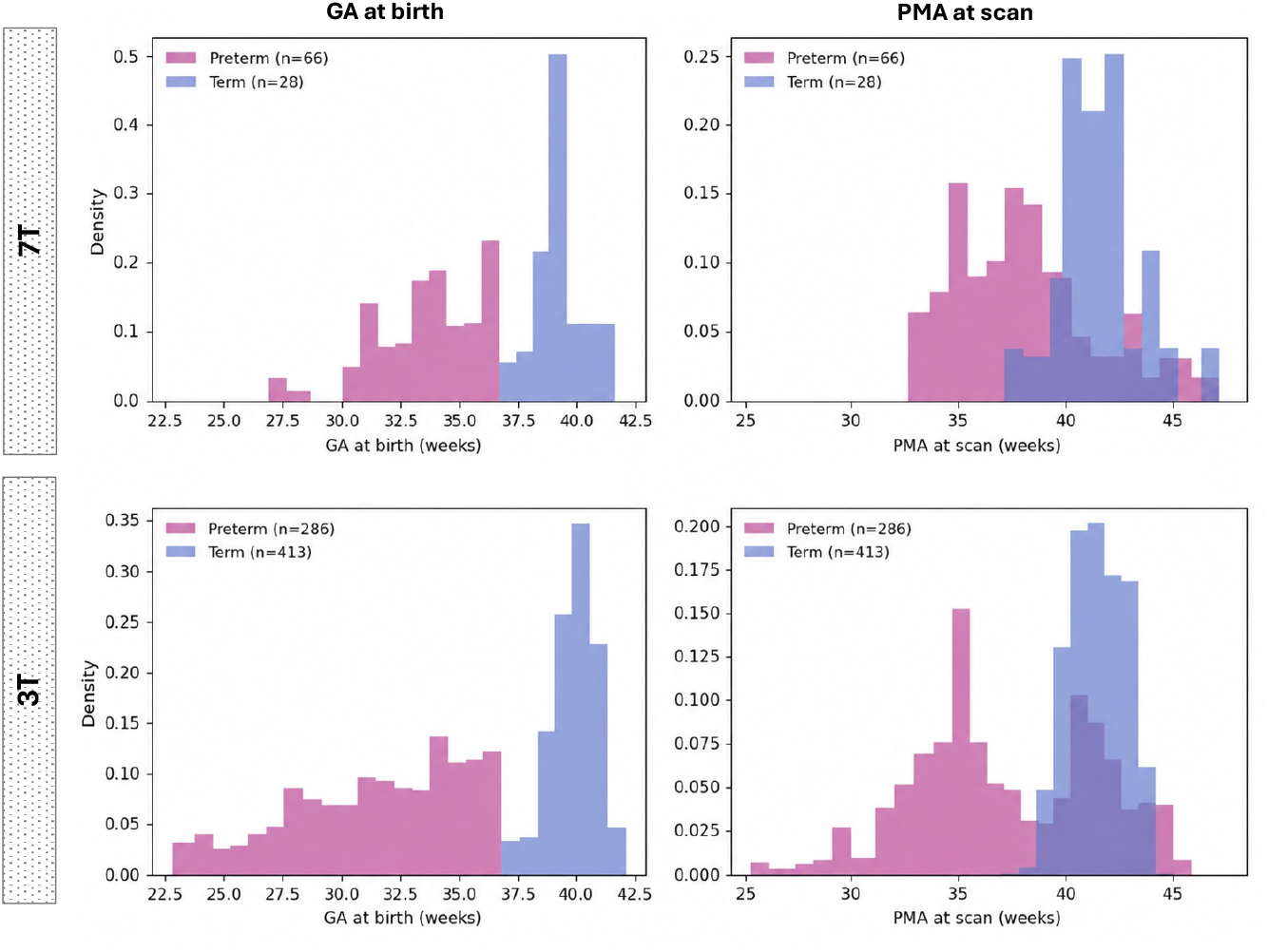
7T and 3T neonatal cohorts used in this work. Histograms show the distribution of GA at birth and PMA at scan in the two cohorts.

**Table 1:** 7T cohort diagnostic classification based on clinical history. Diagnoses are grouped into clinical categories based on primary etiology. Most participants were born preterm or experienced neonatal complications, indicating that the cohort primarily represents infants with early clinical risk factors and suggesting that the segmentation approach was evaluated across a broad range of maturational and anatomical variability. None of the cases were reported to have overt injury of the IC.

| Category | Count | % |
| --- | --- | --- |
| Control / healthy | 12 | 12.8 |
| Preterm / neonatal complications | 59 | 62.8 |
| CHD (congenital heart disease) | 5 | 5.3 |
| Metabolic / maternal conditions | 4 | 4.3 |
| Other / unspecified | 14 | 14.9 |

To assess generalisability of our approach to conventional T2-weighted data, this study also included 3T T2-weighted neonatal brain MRI datasets from the developing Human Connectome Project (dHCP; REC:14/LO/1169). They were acquired on a Philips Achieva system (Philips Healthcare, Netherlands) using a dedicated neonatal imaging protocol (Edwards et al., 2022) (TR/TE = 12000/156 ms, acquired resolution = 0.8 mm, slice thickness = 1.6 mm with 0.8 mm overlap, axial and sagittal planes) and reconstructed to 0.5 mm isotropic resolution (Cordero-Grande et al., 2016). The dataset comprised 413 term (median PMA at scan: 41.57 weeks, range: 37.57-44.86; median PMA at birth: 40.14 weeks, range: 37.00-42.71 ; 218 males) and 286 preterm datasets (median PMA at scan: 36.86 weeks, range: 26.71-45.14; median PMA at birth: 32.29 weeks, range: 23.00-36.86; 155 males) (Fig.1).

All datasets were reviewed by an experienced perinatal neuroradiologist (MR) and radiological evaluation was performed. The 3T term cohort included only the cases without reported significant clinical findings.

### 2.2. Proposed parcellation and assessment protocol

Fig. 2 shows the proposed IC parcellation protocol defined on 3D T2w 7T images. The protocol includes 8 labels: bilateral IC subregions (anterior, posterior, and retro-lenticular limbs) and bilateral PLIC myelination signal ROIs.

**Figure 2:**
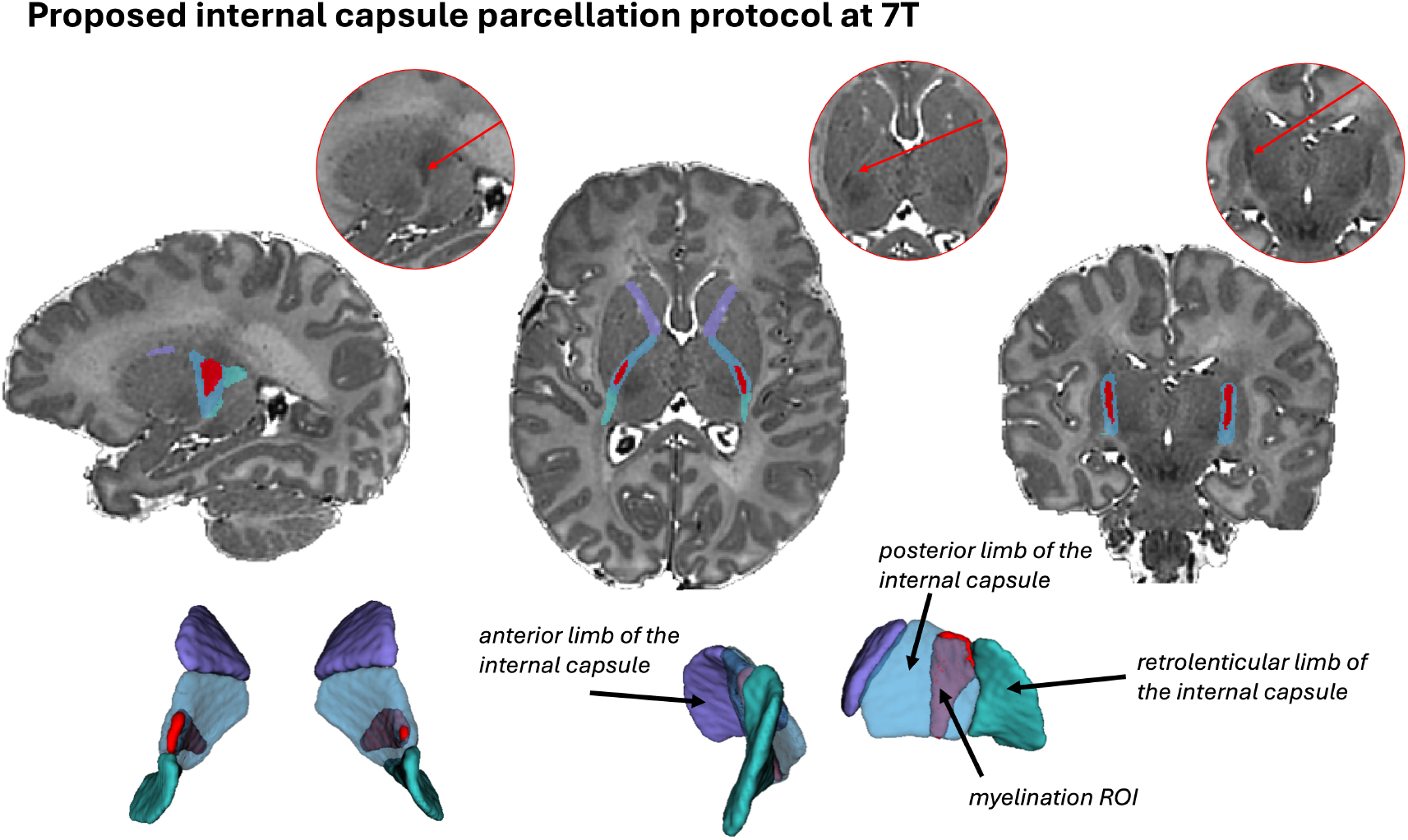
Proposed IC parcellation protocol for T2w neonatal brain MRI at 7T.

Bilateral IC subregion labels were generated through atlas propagation from the neonatal multi-channel MRI atlas (Uus et al., 2021), originally based on definitions in the M-CRIB-WM atlas (Alexander et al., 2020). Bilateral PLIC hypointense signal ROIs, corresponding to regions of T2-weighted contrast associated with myelination, were defined based on conventional perinatal radiological criteria (Arulkumaran) and manually outlined using ITK-SNAP (Yushkevich et al., 2006). The ROIs were visually checked by expert neonatal neuroradiologists (MR, SA).

This protocol allowed extraction of metrics relevant to IC maturation assessment during the neonatal period in terms of both growth and myelination processes. These included: volume of the individual limbs of the IC, volume of the PLIC hypointense ROI, relative hypointense ROI over PLIC volume, and relative hypointense ROI over PLIC signal intensity. For simplicity, the PLIC hypointense ROI is hereafter referred to as the myelination ROI.

### 2.3. Segmentation pipeline

The proposed pipeline for automated IC assessment for 3D T2w neonatal MRI is summarized in Fig. 3.

**Figure 3:**
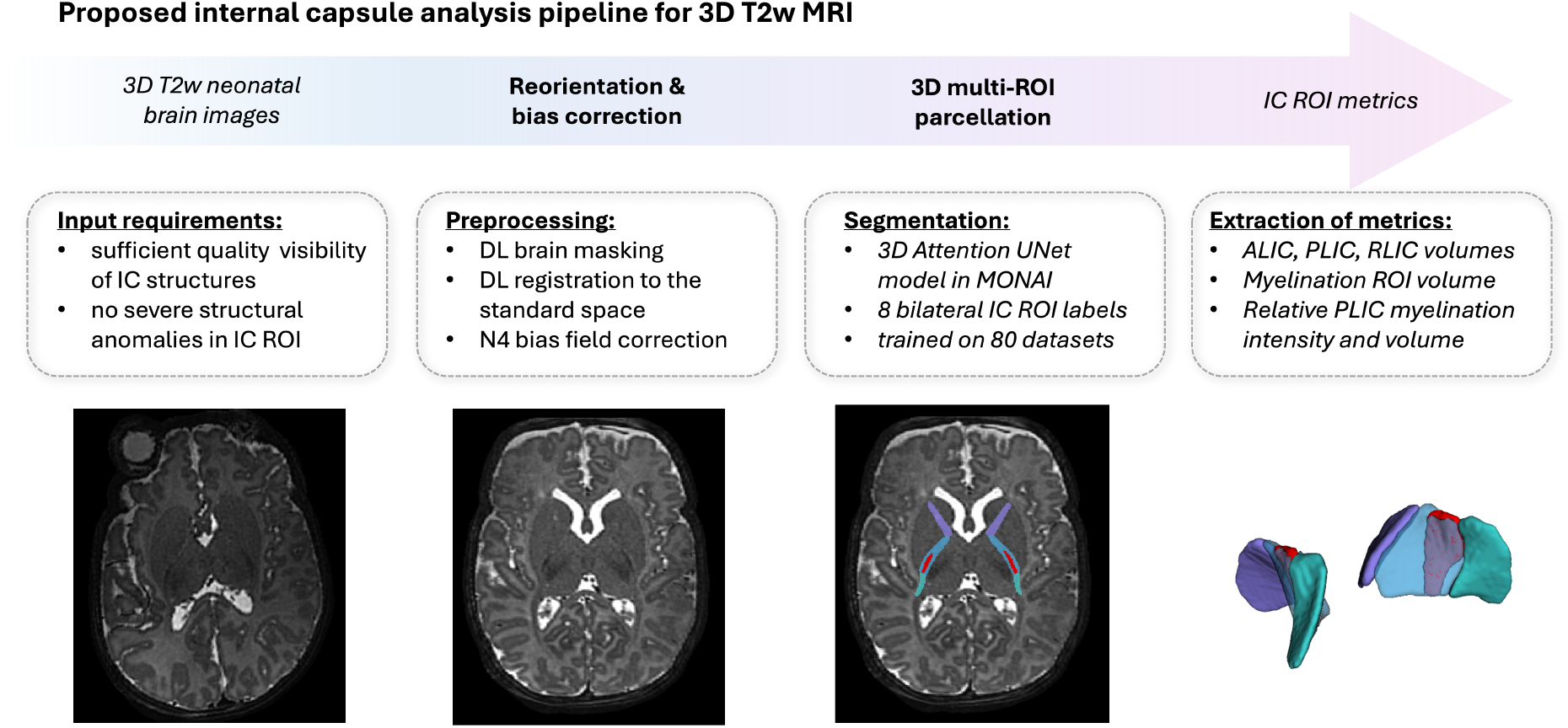
Proposed pipeline for automated segmentation of internal capsule for T2w neontal MRI.

Preprocessing included brain extraction (Uus et al., 2023), spatial reorientation to standard space via hemisphere-based registration (Uus et al., 2023) and N4 bias-field correction (Tustison and Gee, 2009).

Next, IC segmentation was performed using a classical 3D Attention U-Net architecture (Oktay et al., 2018) within the MONAI framework (Cardoso et al., 2022) with five encoder-decoder blocks (output channels 32, 64, 128, 256 and 512), convolution and upsampling kernel size of 3, ReLU activation, dropout ratio of 0.5 and a batch size of 1. We employed AdamW optimiser with a linearly decaying learning rate, initialized at 1×10-4, default beta parameters and weight decay 1×10-5. The training was performed using combined Dice and Cross Entropy Loss with 200,000 iterations. The augmentations included resampling with padding to 256 x 256 x 256 grid, intensity normalisation, random cropping to 128 x 128 x 128 patches, bias field, flipping along sagittal axis and +/-5*◦*rotations. The network was trained on 40 7T and 40 3T datasets specifically selected to include both term and preterm subjects scanner over a 30-44 week scan PMA range. The ground truth labels were created in several stages using the “active learning” strategy by pretraining the preliminary network version on 20 7T datasets with semi-manual propagated labels and running new cases followed by manual refinements in ITK-SNAP and further fine-tuning retraining.

In the final step, the network output is transformed back to the original image space for further extraction of the volumetric and intensity metrics. The code, model weights and executable docker are publicly available online at the following repository: https://github.com/SVRTK/7t-brain-analysis.

The segmentation pipeline was validated on 20 7T and 20 3T independent datasets not used for training. Qualitative assessment was performed by an experienced rater to evaluate anatomical plausibility, boundary accuracy, and preservation of fine structural details. Quantitative agreement with manually refined annotations was assessed using the Dice similarity coefficient and volumetric differences. To assess anatomical localisation of the IC ROIs, the pipeline was also applied to the T2w channel of a multi-channel neonatal brain atlas (Uus et al., 2021), and the resulting labels were mapped onto the corresponding diffusion fractional anisotropy (FA) channel to verify correspondence with the expected IC boundaries.

### 2.4. Quantitative analysis

The proposed pipeline was applied to segment the selected 7T and 3T cohorts. The output IC parcellation labels were used to compute regional volumes (mm^3^), mean normalized T2 intensity within each ROI, and PLIC myelination ratios [myelin ROI volume over PLIC ROI volume]. In addition, to assess normalized intensity changes in PLIC and myelination ROIs, we extracted the average extracerebral cerebrospinal fluid (CSF) intensity from brain tissue parcellations (Uus et al., 2023) for each of the subjects.

#### 2.4.1. 7T cohort

##### Developmental trajectories

Linear mixed-effects models were used to examine age-related changes in regional tissue measures in the 7T sample, adjusting for sex. Two models were evaluated for each outcome: a PMA-only model and a model including both PMA and postnatal age (PNA = PMA − GA ) to assess the influence of pre-maturity at equivalent maturational age. Because some infants contributed multiple scans, a random intercept for infant was included to account for within-subject correlation. For each outcome, linear and quadratic PMA terms were compared using maximum-likelihood estimation (Fisher, 1922) and likelihood ratio testing (Wilks, 1938); the selected fixed-effect structure was refit using Restricted Maximum Likelihood (REML) for final inference (Cook, 2014). Fixed-effect coefficients are reported with 95% confidence intervals. To control for multiple comparisons, false-discovery-rate (FDR) correction using the Benjamini–Hochberg procedure was applied separately within outcome types (IC volumes, PLIC myelin volume ratios, and PLIC myelin intensity ratios) and separately for each predictor (PMA, PNA, and sex) (Benjamini and Hochberg, 1995).

##### Associations with manual biometry

To determine whether automated segmentation outputs reflect established anatomical measurements used in clinical practice, we performed template-guided PLIC biometry in native subject space for the 7T cohort.

A study-specific 7T T2w term template was constructed from reconstructed T2-weighted images using the ANTs multivariate template construction framework (Avants et al., 2011). Template building was performed iteratively using a groupwise registration approach with cross-correlation similarity, multi-resolution optimisation (100×70×50×20 iterations), and gradient step regularization. This yielded an unbiased reference space representative of the cohort.

An expert neuroradiologist (SA) defined landmark pairs in template space for PLIC length, PLIC myelin length, and PLIC myelin thickness, as shown in Fig. 10A. Following non-linear subject-to-template registration, these landmarks were propagated back to each subject’s native image space using the inverse affine and inverse deformation transforms.

For each subject, biometry was quantified as the Euclidean distance between the propagated landmark pairs (i.e. the straight-line distance between the two points in Euclidean space, calculated as the square root of the sum of squared differences between their corresponding coordinates), producing left- and right-sided measures together with a bilateral mean. Associations between template-guided biometry and segmentation-derived metrics were assessed separately in term and preterm infants using Spearman’s rank correlation coefficient (ρ) (Spearman, 1904).

#### 2.4.2. 3T cohort

##### Developmental trajectories

To assess the generalisability of our findings to conventional T2w data, we first examined whether the developmental trajectories observed at 7T were also present in the 3T sample. We characterized age-related variation in IC metrics using sex-adjusted linear regression, as each infant contributed a single scan.

##### Normative modelling and deviation analyses

To capture inter-individual variability and quantify deviations from typical development, we then implemented normative modelling using Gaussian process regression (GPR).^2^ GPR provides both point estimates and predictive uncertainty for each observation, enabling the position of individual infants to be quantified relative to a normative distribution conditional on age and sex. This framework is analogous to paediatric growth charts and allows identification of atypical developmental patterns at the individual level (Dimitrova et al., 2020; O’Muircheartaigh et al., 2020).

We trained GPR models to describe normative IC maturation in the term-born sample using PMA at scan and sex as predictors, and regional tissue metrics as outputs. Separate models were fitted for each metric. The relationship between outcomes and predictors was modelled using a covariance function defined as the sum of a radial basis function kernel, a linear kernel, and a white noise kernel. Model hyperparameters were optimized by maximising the log marginal likelihood. Prediction performance was evaluated using mean absolute error (MAE) between predicted and observed values, estimated using cross-validation within the term-born sample (Dimitrova et al., 2020).

To assess the effects of preterm birth, the model trained on the full term-born dataset was applied to preterm infants scanned at term-equivalent age (n = 134). A Z-score was derived for each individual in both the preterm and term-born samples by computing the difference between the observed value and the model prediction, normalized by the predictive uncertainty (i.e. the square root of the predictive variance).

We then investigated the association between prematurity and IC metrics by (1) calculating correlations with GA at birth (Spearman’s ρ) and (2) testing group differences between preterm and term infants in signed Z-scores using the Mann–Whitney U test and Vargha–Delaney A effect size (Vargha and Delaney, 2000; Dimitrova et al., 2020). Multiple comparison correction was performed using the Benjamini–Hochberg procedure, applied separately within outcome types (IC volumes, PLIC myelin volume ratios, and PLIC myelin intensity ratios) (Benjamini and Hochberg, 1995).

## 3. Results

### 3.1. Segmentation performance

The proposed segmentation pipeline showed strong anatomical correspondence with internal capsule boundaries on the atlas FA map (Fig. 4), supporting the anatomical validity of the T2w-driven parcellation approach. Evaluation results on 40 independent 3T and 7T datasets compared with ground-truth manual refinement of the network outputs are summarized in Tab. 2. The pipeline delineated IC subregions with generally acceptable anatomical accuracy, preserving small structures and myelination-related signal boundaries within the PLIC.

**Table 2:** Evaluation of the segmentation pipeline vs. manually refined labels.

| Metric | ALIC | PLIC | RLIC | Myelination ROI |
| --- | --- | --- | --- | --- |
| 20 7T datasets: |  |  |  |  |
| Relative volume difference | 1.44% ± 1.30% | 1.46% ± 1.72% | 1.97% ± 1.76% | 2.93% ± 2.68% |
| Dice | 0.983 ± 0.022 | 0.980 ± 0.019 | 0.970 ± 0.019 | 0.971 ± 0.019 |
| 20 3T datasets: |  |  |  |  |
| Relative volume difference | 1.66% ± 1.52% | 1.97% ± 2.49% | 2.64% ± 2.13% | 4.80% ± 3.78% |
| Dice | 0.973 ± 0.026 | 0.966 ± 0.031 | 0.947 ± 0.031 | 0.950 ± 0.031 |

**Figure 4:**
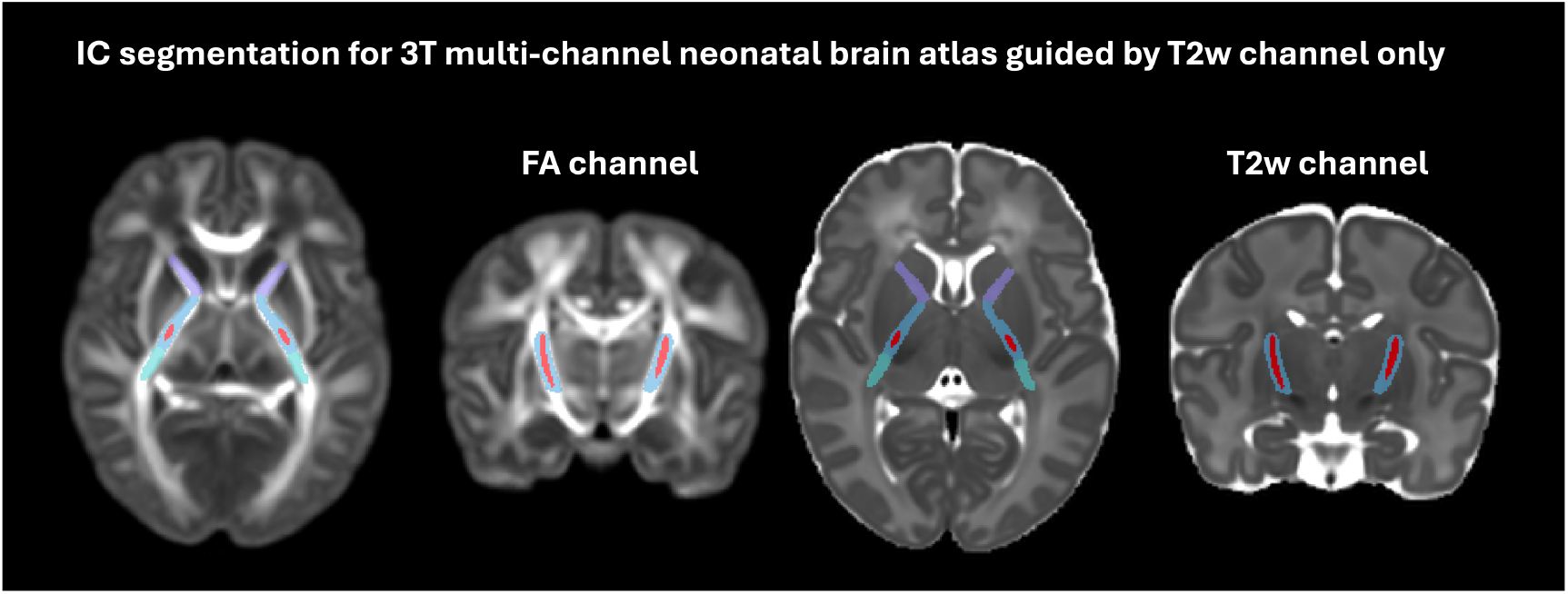
Example of the IC segmentation output generated from the T2w channel of the atlas overlayed over the FA channel of the atlas (Uus et al., 2021).

Segmentation quality varied across subjects, influenced by factors such as local SNR, 3-D reconstruction quality, low signal contrast due to prematurity and the presence of blood vessels adjacent to the myelination ROI. The average Dice scores were > 0.95 for all ROIs and average relative volume differences were < 5%. Differences in the relative myelination ROI over PLIC intensity ratio were below 1% (7T: 0.47% ± 0.47%; 3T: 0.45% ± 0.38%). Accuracy was consistently higher at 7T than at 3T, in keeping with the superior tissue contrast and structural visibility of the 7T images. Manual inspection and refinement required less than 2 minutes per case on average.

### 3.2. Quantitative analysis

#### 3.2.1. 7T cohort

##### Developmental trajectories

Tab. 3 and Fig. 5 summarize the quantitative analysis results for the 7T cohort. An example of processed longitudinal dataset in shown in Fig. 6.

**Table 3:** 7T mixed-effects (REML) model coefficients per week. Raw p-values and Benjamini–Hochberg FDR-corrected q-values are shown. FDR correction was applied separately within outcome families (absolute volumes; relative intensity vs CSF; relative myelin vs PLIC) for each predictor (PMA, PNA, sex).

| Region / Metric | $\beta_{PMA}$ (95% CI) | p(PMA) | q(PMA) | $\beta_{PNA}$ (95% CI) | p(PNA) | q(PNA) | $\beta_{Sex}$ (95% CI) | p(Sex) | q(Sex) |
| --- | --- | --- | --- | --- | --- | --- | --- | --- | --- |
| Absolute ALIC volume | 0.0189 [0.0151, 0.0227] | <0.001 | <0.001 | 0.0014 [-0.0037, 0.0065] | 0.594 | 0.650 | 0.0152 [-0.0079, 0.0384] | 0.198 | 0.264 |
| Absolute PLIC volume | 0.0250 [0.0208, 0.0291] | <0.001 | <0.001 | 0.0009 [-0.0030, 0.0048] | 0.650 | 0.650 | 0.0359 [0.0109, 0.0608] | 0.005 | 0.020 |
| Absolute RLIC volume | 0.0152 [0.0125, 0.0179] | <0.001 | <0.001 | -0.0008 [-0.0033, 0.0016] | 0.502 | 0.650 | 0.0185 [0.0028, 0.0341] | 0.020 | 0.040 |
| Absolute PLIC myelin volume | 0.0110 [0.0098, 0.0122] | <0.001 | <0.001 | 0.0012 [0.0001, 0.0024] | 0.038 | 0.152 | 0.0012 [-0.0065, 0.0088] | 0.768 | 0.768 |
| Relative PLIC/CSF intensity | -0.0135 [-0.0154, -0.0115] | <0.001 | <0.001 | 0.0005 [-0.0014, 0.0023] | 0.622 | 0.622 | 0.0029 [-0.0074, 0.0132] | 0.585 | 0.585 |
| Relative myelin/CSF intensity | -0.0161 [-0.0179, -0.0143] | <0.001 | <0.001 | 0.0013 [-0.0005, 0.0030] | 0.158 | 0.237 | 0.0047 [-0.0053, 0.0146] | 0.359 | 0.576 |
| Relative myelin/PLIC intensity | -0.0140 [-0.0161, -0.0118] | <0.001 | <0.001 | 0.0033 [0.0013, 0.0053] | 0.001 | 0.003 | 0.0057 [-0.0071, 0.0185] | 0.384 | 0.576 |
| Relative myelin/PLIC volume | 0.0133 [0.0113, 0.0153] | <0.001 | <0.001 | 0.0008 [-0.0010, 0.0027] | 0.380 | 0.380 | -0.0081 [-0.0200, 0.0038] | 0.183 | 0.183 |

**Figure 5:**
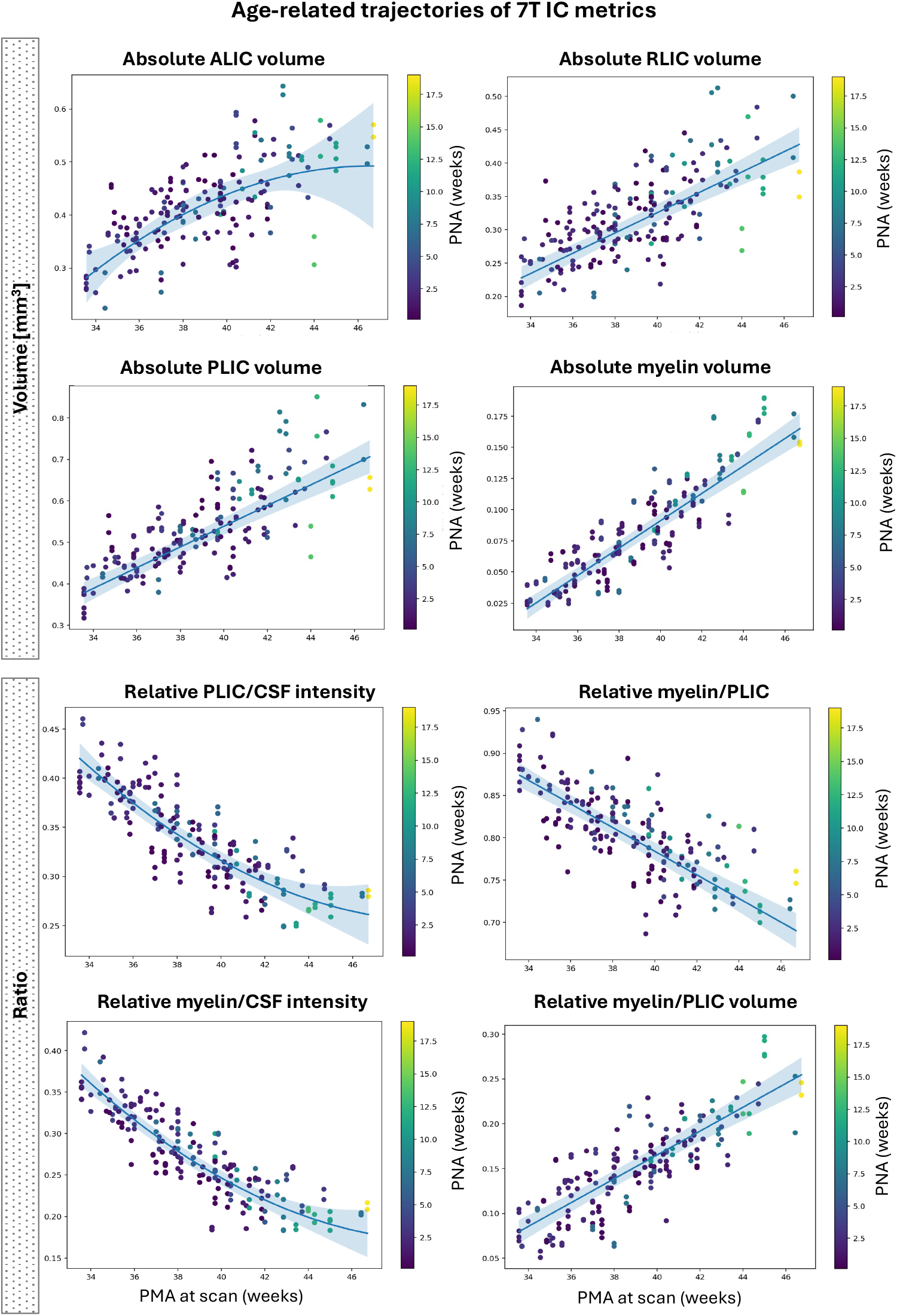
Age-related trajectories of 7T IC myelin metrics. Observed values are plotted against PMA at scan, with points coloured by PNA. Solid lines show the model-selected (linear vs quadratic by AIC) population-level mixed-effects fit (random intercept for infant); shaded bands indicate 95% confidence intervals for the fixed-effects mean.

**Figure 6:**
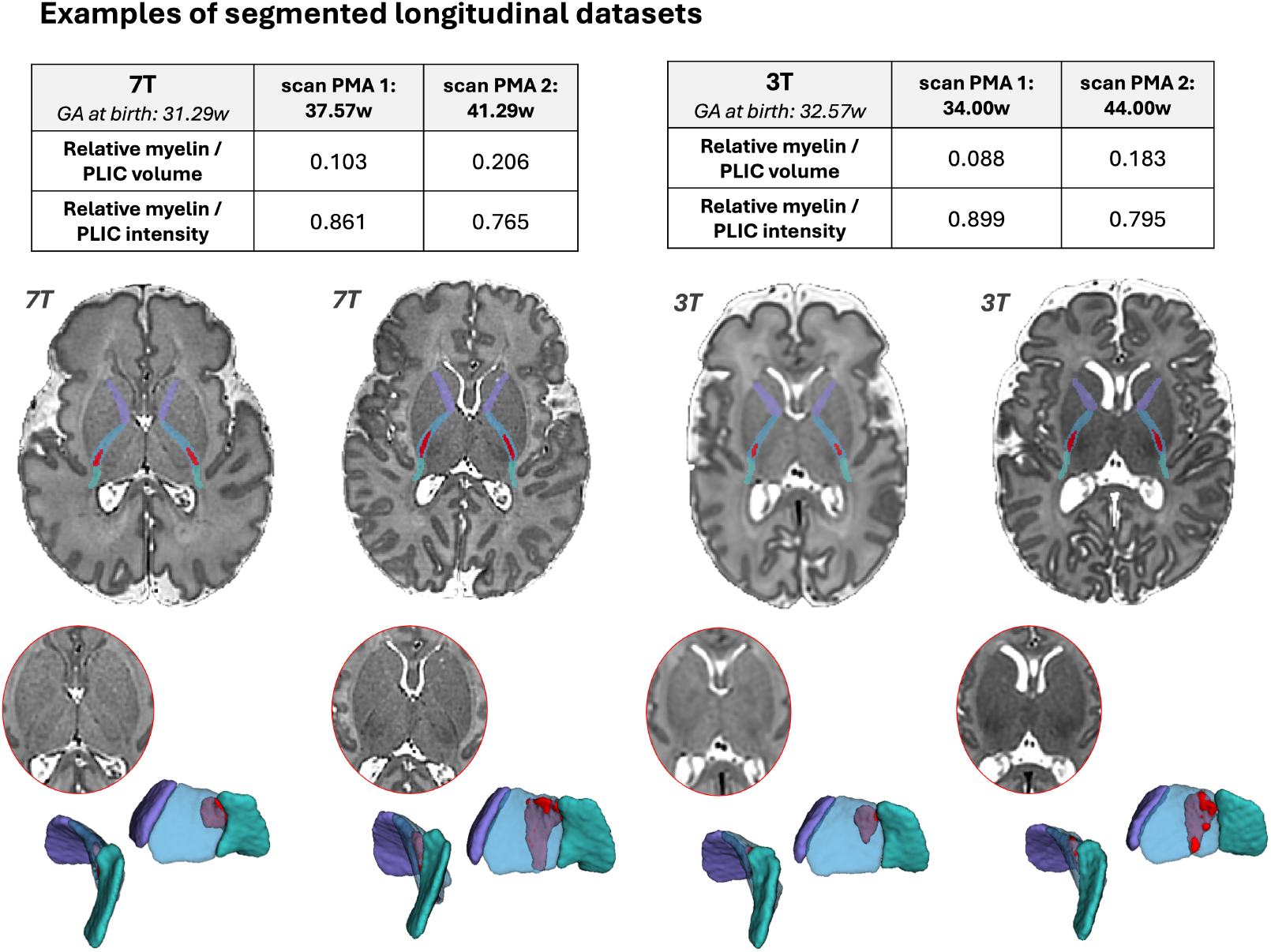
Examples of processed longitudinal datasets at 7T and 3T. Automated 3D segmentation of the IC is shown at two timepoints in the same infant. Between scans, the relative PLIC myelination volume increased, while mean-normalized T2 intensity decreased, consistent with progressive myelination and maturation of the IC.

Mixed-effects models demonstrated robust associations between PMA and all structural metrics after FDR correction. All absolute volume measures showed significant positive associations with PMA (all q < 0.001). Quadratic PMA effects remained significant for ALIC and RLIC volumes after FDR correction, consistent with non-linear maturation effects across the sampled age range.

Relative intensity and ratio metrics also showed strong PMA associations. Both PLIC/CSF and myelin/CSF intensity ratios showed significant negative associations with PMA after correction (all q < 0.001), indicating decreases across the scanned age range. Relative myelin/PLIC intensity and relative myelin/PLIC volume were likewise significantly associated with PMA following FDR adjustment.

When PNA was added to the model, only the relative myelin/PLIC intensity measure remained significant after correction (q = 0.003). At equivalent PMA, higher PNA was associated with a higher myelin-to-PLIC intensity ratio, indicating reduced relative hypointensity within the myelinated PLIC compared to the non-myelinated portion. No volumetric measure showed a significant association with PNA after FDR correction.

For sex, absolute PLIC and RLIC volumes remained significant after FDR adjustment (PLIC: q = 0.020; RLIC: q = 0.040), while all other sex effects were attenuated following correction.

##### Associations with manual biometry

Fig. 10 shows associations between biometry and segmentation derived measures in the 7T cohort.

Across both term and preterm infants, PLIC length, myelin segment length, and myelin thickness showed consistent positive associations with absolute PLIC volume and absolute PLIC myelin volume (all ρ 0.4–0.8, p < 0.05), indicating that larger manually measured tract dimensions correspond to larger segmented volumes. Similar positive relationships were observed for the relative myelin/PLIC volume metric, although these were generally weaker.

Associations between biometry and intensity-based metrics differed by group and by the specific biometry measure. For PLIC length, correlations with intensity ratios were negative and moderate-to-strong in preterm infants, but weak or near-zero in term-born infants. For myelin length and myelin thickness, negative correlations were observed more consistently across groups, although effect sizes and value distributions differed between term and preterm infants. In addition, term-born infants showed a shift towards greater myelin thickness alongside lower intensity ratio values, consistent with more advanced maturation and a reduced dynamic range in T2w contrast.

#### 3.2.2. 3T cohort

##### Developmental trajectories

Tab. 4 and Fig. 7 the developmental trajectory analyses in the 3T cohort. In this larger cross-sectional sample, PMA showed significant associations with all volumetric and relative intensity measures after FDR correction. All absolute volume metrics increased with PMA, whereas relative PLIC/CSF, relative myelin/CSF, and relative myelin/PLIC intensity ratios decreased with PMA. Relative myelin/PLIC volume increased with PMA. These age-related patterns were directionally consistent with those observed in the 7T cohort.

**Table 4:**
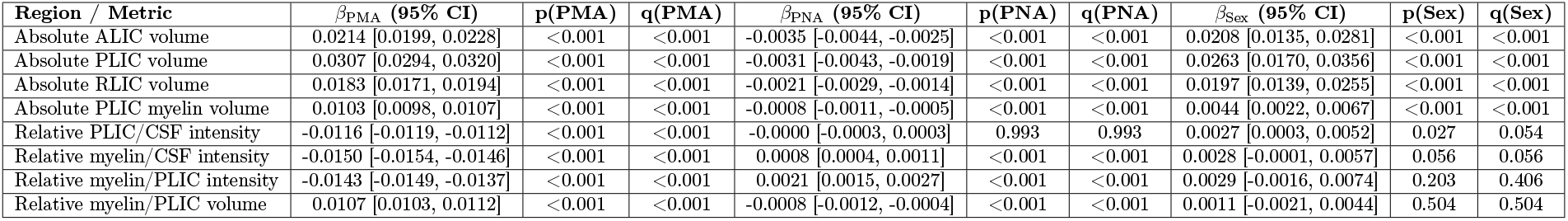
3T regression model coefficients per week. Raw p-values and Benjamini–Hochberg FDR-corrected q-values are shown. FDR correction was applied separately within outcome families (absolute volumes; relative intensity vs CSF; relative myelin vs PLIC) for each predictor (PMA, PNA, sex).

**Figure 7:**
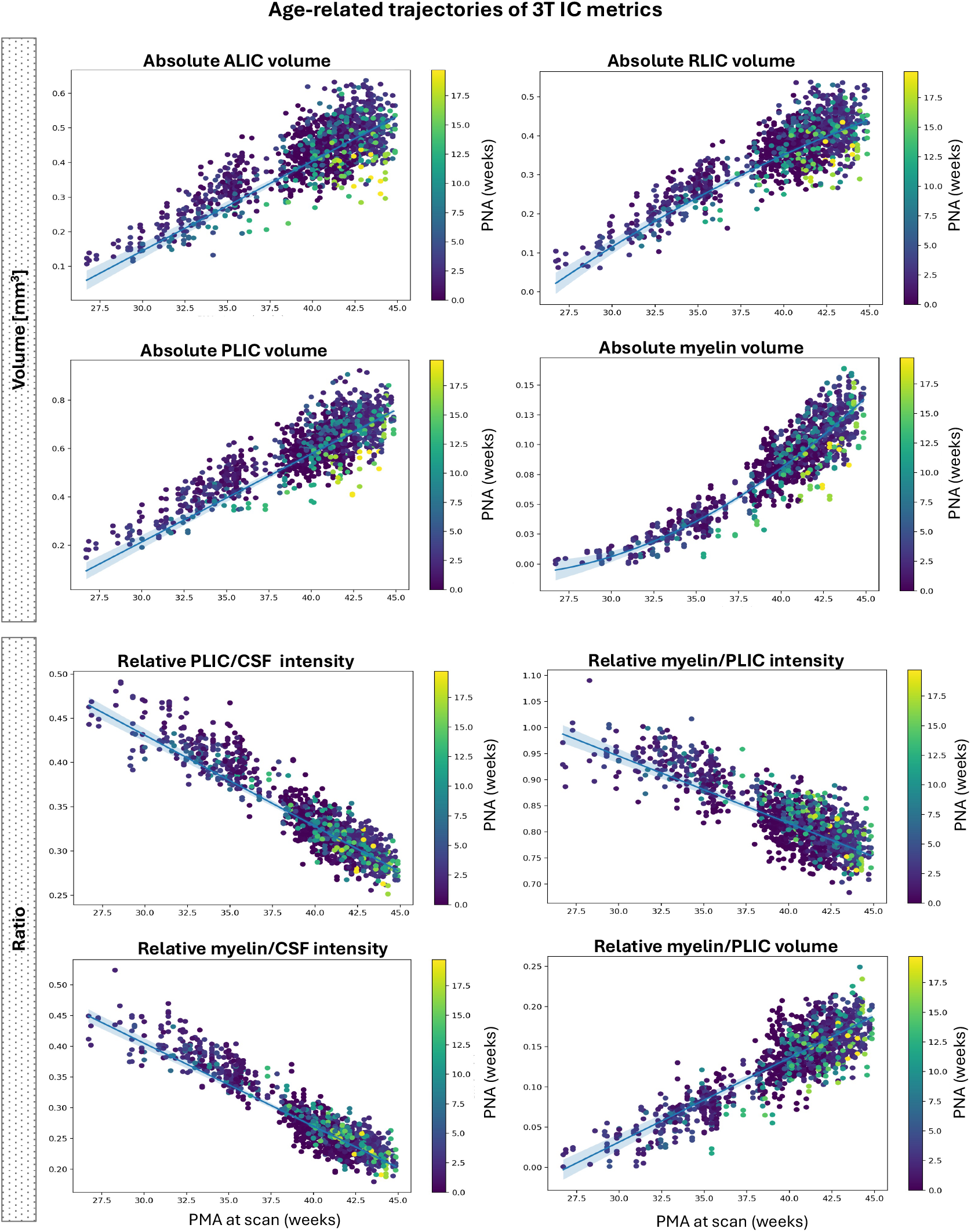
Age-related trajectories of 3T IC metrics. Observed values are plotted against PMA at scan, with points coloured by PNA. For each outcome, linear versus quadratic PMA terms were compared, and the selected model is shown. Solid lines show the fitted population-level regression curve (evaluated at the median PNA); shaded bands indicate 95% confidence intervals for the mean prediction. Bottom: Linear regression models. Raw p-values and FDR-corrected q-values are reported. FDR correction was applied separately within outcome families (absolute volumes; relative intensity vs CSF; relative myelin vs PLIC) and separately for each predictor (PMA, PNA, sex).

PNA effects were also prominent at 3T. After FDR correction, higher PNA at equivalent PMA was associated with lower absolute volumes of the ALIC, PLIC, and RLIC, as well as reduced PLIC myelin volume. In contrast, higher PNA was associated with increased relative intensity measures, including both the myelinated PLIC/CSF and myelinated PLIC/unmyelinated PLIC intensity ratios, and with a reduced myelinated PLIC/unmyelinated PLIC volume ratio. No association was observed for the unmyelinated PLIC/CSF intensity ratio.

##### Normative modelling and deviation analyses

As shown in Tab. 5, normative modelling demonstrated moderate-to-good predictive performance across all metrics, with cross-validated Spearman correlations ranging from 0.45 to 0.75. The strongest prediction was observed for absolute PLIC volume and relative PLIC/CSF intensity, followed by PLIC myelin volume and relative myelin/CSF intensity. Fig. 8 illustrates the normative trajectories and the distribution of preterm infants around them.

**Table 5:** Signed normative deviation (Z-score) analyses across internal capsule metrics. Z-scores represent deviation from the normative model adjusted for PMA at scan and sex. Associations with gestational age (GA) at birth were assessed within the preterm group using Spearman correlation. Group differences were assessed using Mann–Whitney U tests. False-discovery-rate (FDR) correction (Benjamini–Hochberg) was applied separately within outcome families and separately for each analysis.

| Region / Metric | Spearman $\rho$ (CV) | Term mean Z | Preterm mean Z | GA vs Z $\rho$ | $p$ | FDR $q$ | Mann–Whitney U | $p$ | FDR $q$ | A |
| --- | --- | --- | --- | --- | --- | --- | --- | --- | --- | --- |
| Absolute ALIC volume | 0.58 | 0.01 | -0.35 | 0.492 | $1.51 \times 10^{-9}$ | $2.01 \times 10^{-9}$ | 23714 | 0.0128 | 0.0513 | 0.43 |
| Absolute PLIC volume | 0.75 | 0.01 | -0.04 | 0.583 | $1.42 \times 10^{-13}$ | $5.67 \times 10^{-13}$ | 28114 | 0.7807 | 0.7807 | 0.51 |
| Absolute RLIC volume | 0.67 | 0.00 | -0.25 | 0.527 | $6.08 \times 10^{-11}$ | $1.22 \times 10^{-10}$ | 24647 | 0.0572 | 0.1144 | 0.45 |
| Absolute PLIC myelin volume | 0.73 | 0.00 | -0.20 | 0.477 | $5.74 \times 10^{-9}$ | $5.74 \times 10^{-9}$ | 26008 | 0.2957 | 0.3942 | 0.47 |
| Relative PLIC/CSF intensity | 0.75 | 0.00 | 0.16 | 0.148 | 0.0885 | 0.1327 | 29864 | 0.1678 | 0.1678 | 0.54 |
| Relative myelin/CSF intensity | 0.72 | 0.00 | 0.44 | 0.032 | 0.7109 | 0.7109 | 34518 | $1.66 \times 10^{-5}$ | $2.49 \times 10^{-5}$ | 0.62 |
| Relative myelin/PLIC intensity | 0.45 | 0.01 | 0.58 | -0.196 | 0.0231 | 0.0693 | 36807 | $9.11 \times 10^{-9}$ | $2.73 \times 10^{-8}$ | 0.67 |
| Relative myelin/PLIC volume | 0.52 | 0.00 | -0.26 | 0.106 | 0.2248 | 0.2248 | 24005 | 0.0211 | 0.0211 | 0.43 |

**Figure 8:**
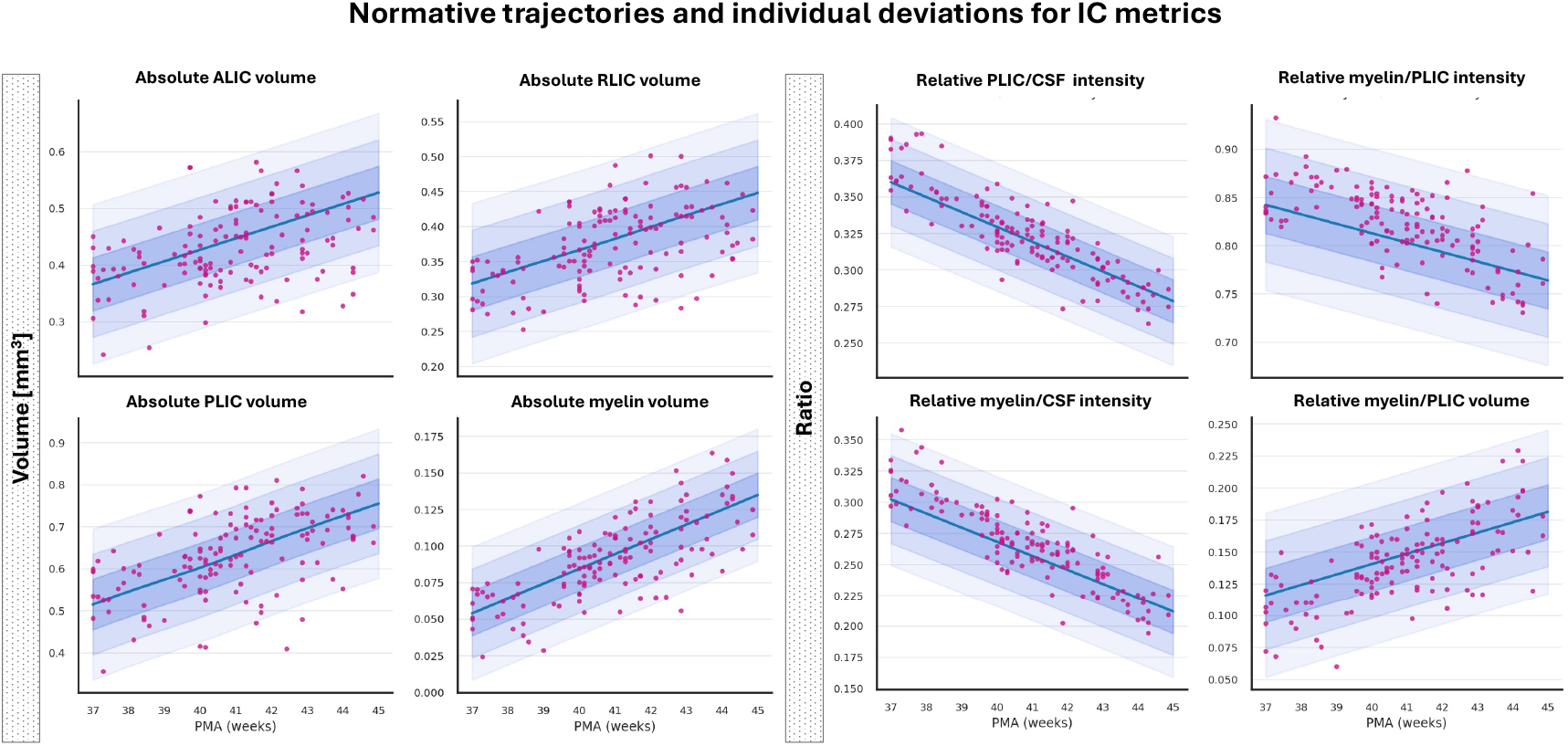
Normative trajectories and individual deviations for IC metrics. Normative developmental trajectories were estimated in the term-born reference sample. For each metric, the solid line represents the mean predicted value as a function of PMA, with shaded regions indicating ±1, ±2, and ±3 standard deviations of the model uncertainty. Individual preterm infants scanned at term-equivalent age are shown in magenta. All trajectories are adjusted for sex. Deviations of preterm infants from the normative distribution are visually apparent particularly for relative intensity measures.

Within the preterm group scanned at term-equivalent age, GA at birth was significantly associated with Z-scores for all volumetric metrics after FDR correction, including ALIC volume (ρ = 0.49, q < 0.001), PLIC volume (ρ = 0.58, q < 0.001), RLIC volume (ρ = 0.53, q < 0.001), and PLIC myelin volume (ρ = 0.48, q < 0.001). No intensity or ratio metric showed a significant association with GA at birth after correction.

In contrast, group comparisons between preterm and term infants showed significant group differences only for myelin-related metrics. Preterm infants showed higher signed Z-scores for relative myelin intensity versus CSF (mean Z = 0.44 vs 0.00; q < 0.001), relative myelin versus PLIC intensity (mean Z = 0.58 vs 0.01; q < 0.001), and the myelin-to-PLIC volume ratio (mean Z = -0.26 vs 0.00; q = 0.02). No absolute volume metric survived FDR correction in the group comparison, although ALIC volume showed a borderline effect (q = 0.051). Fig. 9 and Tab. 5 summarise these deviation analyses.

**Figure 9:**
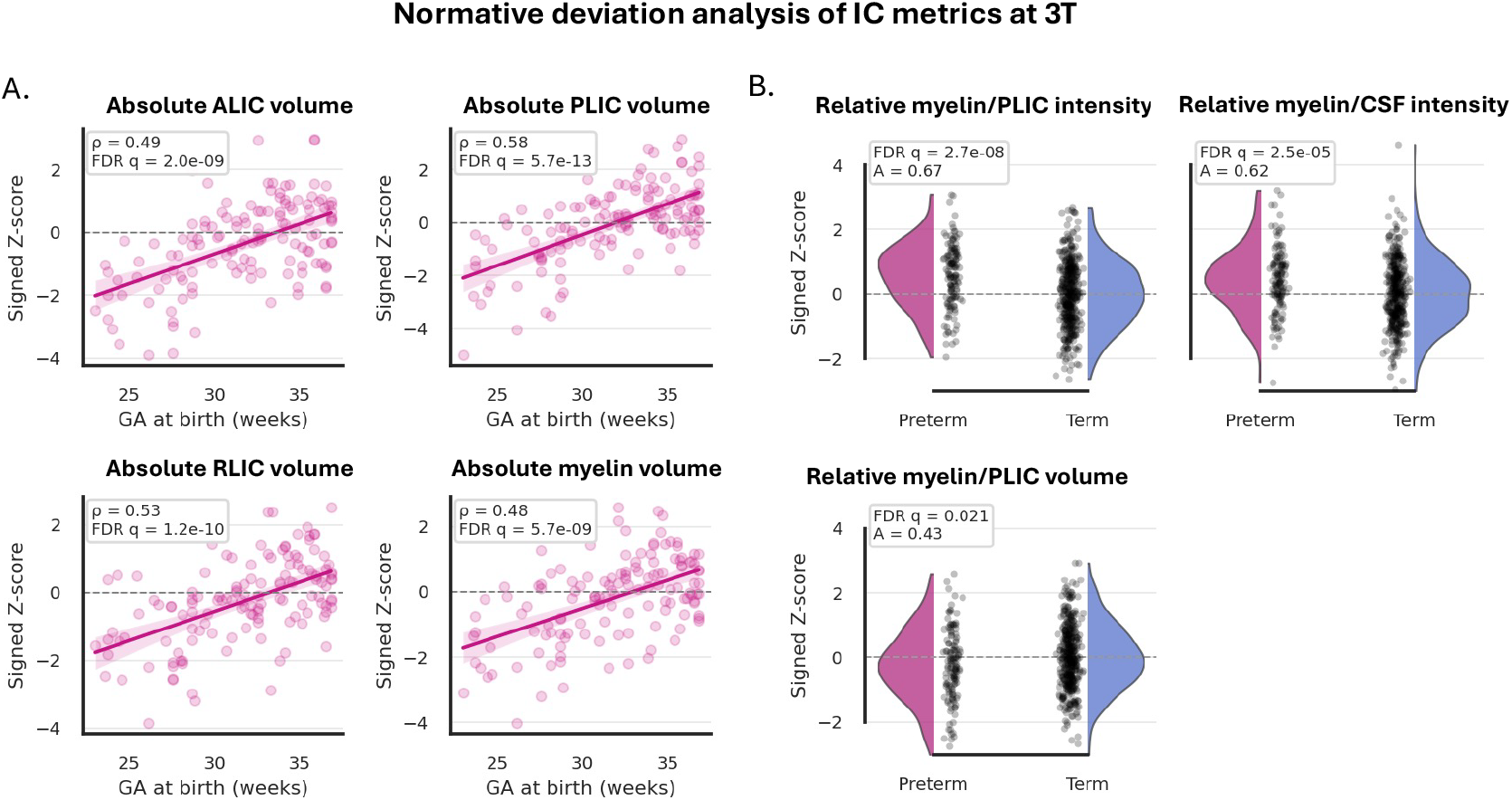
Normative deviation analysis of IC metrics. (A) Significant associations between GA at birth and Z-scores within the preterm group. (B) Significant group differences between preterm infants scanned at term-equivalent age and term-born controls

**Figure 10:**
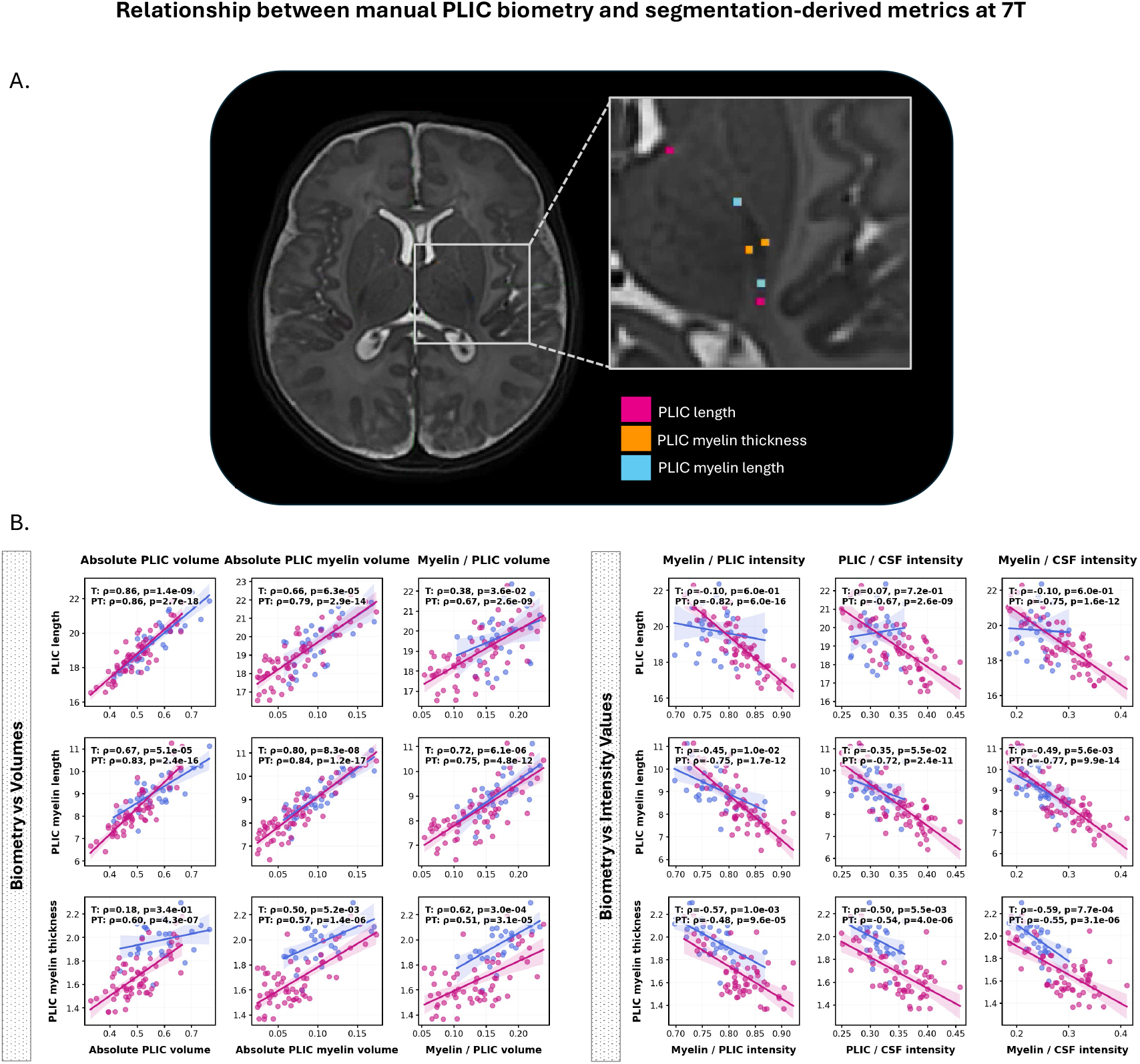
Relationship between manual PLIC biometry and segmentation-derived metrics at 7T.(A) Illustration of manual radiological biometry, showing measurement of PLIC length, myelin thickness, and myelin length. (B) Associations between manual biometry measures and segmentation-derived metrics. Left panels show relationships with volumetric measures, while right panels show relationships with intensity-based measures.

## 4. Discussion

In this work, we present a fully automated framework for anatomically detailed segmentation and quantitative assessment of the neonatal IC using structural T2w MRI. We further evaluate its feasibility and potential relevance for large-scale research studies and clinical applications.

Leveraging the enhanced spatial resolution and tissue contrast of 7T imaging, we defined a multi-regional 3D parcellation protocol comprising eight labels, including the anterior, posterior, and retrolenticular limbs of the IC in each hemisphere, together with a bilateral PLIC myelination ROI. This protocol enabled training of a dedicated deep learning pipeline for rapid segmentation and extraction of volumetric and relative intensity metrics of IC maturation.

To support translation to the clinical setting, the model was trained using both high-resolution 7T neonatal T2-weighted scans (n = 40) and conventional 3T scans (n = 40). Evaluation in 40 independent datasets showed robust performance across both field strengths, with Dice scores exceeding > 0.95 for all ROIs and relative volume differences below < 5%, indicating accurate delimitation of IC subregions and myelination-related signal boundaries.

### 4.1. Developmental trajectories

Across both 7T and 3T cohorts, IC metrics showed robust and consistent associations with PMA at scan. Specifically, absolute volumetric measures increased with age, while relative intensity measures decreased, reflecting progressive white matter maturation. These findings are consistent with the known trajectory of early myelination, which is associated with progressive changes in tissue composition, including increasing myelin content and reductions in water fraction (Deoni et al., 2012). In structural MRI, these changes are reflected in reductions in T2w signal intensity with respect to specific PMA period(Counsell et al., 2002a).

A key finding of this study is that volumetric and intensity-derived measures showed distinct sensitivity to neonatal IC maturation. Volumetric metrics exhibited highly consistent age-related associations across cohorts, indicating that they provide stable markers of developmental change in IC structure. In contrast, intensity-based measures appeared more sensitive to early maturational variation and showed stronger associations with prematurity-related factors, particularly in the larger 3T cohort.

This difference likely reflects the underlying biology of early white matter maturation. During the preterm period, T2w signal remains highly sensitive to microstructural tissue change, including variation in water content, tissue organization, and myelination-related processes. As a result, intensity-based measures may retain greater sensitivity to early developmental variation and prematurity-related deviation. By term-equivalent age, when the PLIC is more mature and T2w contrast is reduced, this sensitivity may be attenuated, while volumetric measures continue to provide a more consistent index of structural maturation.

### 4.2. Effects of prematurity and postnatal exposure

By modelling PNA alongside PMA, we were able to separate maturational effects from variation related to postnatal exposure at equivalent developmental stage. At 7T, PNA effects were relatively subtle and only observed in the relative myelination ROI/PLIC intensity metric potentially due to the small number of cases. In contrast, in the larger 3T cohort, PNA effects were robust across both volumetric and intensity-based measures.

At equivalent PMA, higher PNA, corresponding to lower gestational age at birth, was associated with reduced myelination-related contrast and differences in volumetric measures. This suggests that these effects reflect prematurity-related modulation of white matter development rather than maturation alone. These findings support the interpretation that variation in postnatal extrauterine exposure contributes to early white matter development, consistent with prior work demonstrating altered microstructural maturation in preterm infants (Dimitrova et al., 2020; O’Muircheartaigh et al., 2020).

### 4.3. Relationship to manual biometry

Associations between segmentation-derived metrics and manual PLIC biometry support the biological validity of the proposed measures. Volumetric metrics showed positive relationships with manually defined tract dimensions, with greater PLIC length, myelin segment length, and myelin thickness corresponding to larger segmented PLIC and myelination ROI volumes.

In contrast, intensity-based metrics showed negative associations with manual biometry, with greater tract dimensions corresponding to lower T2-weighted signal, consistent with increased myelination accompanying tract growth and maturation.

Importantly, these relationships differed by both group and biometry measure. Associations between PLIC length and intensity-based metrics were evident primarily in preterm infants, whereas term-born infants showed weak or absent relationships. For myelin length and myelin thickness, negative associations were observed more consistently across groups, although effect sizes were attenuated in term-born infants. In addition, term-born infants showed a shift towards greater myelin thickness alongside lower intensity ratio values, consistent with more advanced maturation and a reduced dynamic range in T2-weighted contrast.

Together, these findings indicate that volumetric and intensity-derived measures capture complementary but distinct aspects of white matter maturation, with volumetric measures reflecting macroscopic structural growth and intensity-derived metrics showing greater sensitivity to early developmental variation.

### 4.4. Generalizability and normative modelling

A key strength of this work is the demonstration that IC metrics derived from 7T structural imaging generalize to conventional 3T data. Despite differences in image quality and contrast, the same developmental patterns were observed, supporting the robustness and translational potential of the approach.

Normative modelling further extends these findings by enabling individual-level inference. By defining typical developmental trajectories in term-born infants, we show that preterm infants scanned at term-equivalent age exhibit systematic deviations, particularly in myelination-related intensity and ratio measures. Although volumetric metrics showed strong associations with gestational age at birth, group-level deviations at term-equivalent age were primarily captured by intensity-based measures.

Importantly, this pattern is consistent with the PNA-based analyses, suggesting that prematurity-related effects observed at equivalent PMA reflect altered developmental trajectories rather than simple maturational delay. Together, these findings highlight that intensity-based measures are particularly sensitive to early developmental deviation associated with preterm birth.

### 4.5. Methodological implications

This work demonstrates that structural T2-weighted MRI, when combined with anatomically informed segmentation, can yield quantitative markers of white matter maturation without the need for specialized quantitative MRI acquisitions. This is particularly important in clinical and large-scale research settings, where acquisition time, protocol complexity, and processing burden are often constrained. More broadly, the integration of multi-regional parcellation, automated segmentation, and normative modelling provides a scalable, objective, and anatomically informed framework for assessing regional IC maturation across field strengths and populations, while complementing conventional qualitative radiological evaluation.

To facilitate the clinical translation and reproducibility of our findings, the source code, trained model weights, and an executable Docker container are publicly available at https://github.com/SVRTK/7t-brain-analysis.

### 4.6. Limitations and future directions

While our framework demonstrates high technical accuracy and biological validity, we acknowledge several limitations that provide a roadmap for future development.

Firstly, the current pipeline was not extensively trained or evaluated across the full range of confirmed severe PLIC abnormalities or deep grey matter anomalies. Extending the framework to these pathological cases, together with further formalization of the normative modelling approach, will be a critically important next step towards establishing its clinical utility for anomaly detection and characterisation. For example, in hypoxic–ischaemic encephalopathy, the PLIC myelination ROI may exhibit both reduced and increased T2-weighted signal, as well as altered morphology.

Additionally, automated quality control of both input image quality and segmentation accuracy will be essential for reliable application in large, heterogeneous cohorts. In this context, combining atlas-based label propagation with the 3D Attention U-Net framework may provide a robust strategy to mitigate suboptimal performance in challenging cases. Further extension and validation across additional acquisition protocols and field strengths, including 1.5T, will also be required.

A further limitation relates to the interpretation of volumetric measures of the IC. Although the IC can be consistently delineated using anatomically informed segmentation, its boundaries on T2w imaging are not always sharply defined, particularly in early developmental stages where tissue contrast remains low. As a result, the absolute volume of IC subregions is better interpreted as an operational measure that depends on the segmentation protocol rather than a direct reflection of a discrete anatomical structure. Importantly, because the same segmentation framework is applied uniformly across all subjects, any systematic over- or underestimation is expected to be consistent across the cohort, allowing for robust relative comparisons and the assessment of developmental trends. While the strong and consistent associations observed with age support the utility of these measures as markers of structural maturation, caution is warranted when attributing biological specificity to absolute volumetric values.

Finally, a central tenet of this study is the use of T2w signal intensity as a marker for the progression of myelination. While we ensured the anatomical validity of this structural-only approach by mapping our T2w-driven labels onto a neonatal FA atlas, confirming strong correspondence with the expected IC boundaries, T2w intensity remains an indirect marker of the underlying maturation process. Consequently, signal changes may be influenced by factors beyond myelination, such as changes in water content or tissue density. Future work will focus on integrating T1w imaging, quantitative T1, T2, and T2* contrasts, diffusion MRI, and susceptibility mapping (QSM) to improve sensitivity to specific microstructural tissue properties (Deoni et al., 2012). In parallel, our parcellation protocol will be further extended and refined to include additional deep grey matter structures, allowing for a more comprehensive assessment of neonatal brain development.

## 5. Conclusions

We present the first 3D deep learning pipeline for automated segmentation and quantitative assessment of the neonatal internal capsule (IC) and PLIC myelination on 7T and 3T T2-weighted structural MRI.

The approach enables detailed parcellation of the three internal capsule limbs in each hemisphere together with the PLIC myelination ROI, followed by objective quantification of regional volume and signal intensity. In doing so, it provides a structural MRI-based framework for characterising early myelination in vivo. This work establishes a foundation for the development of robust quantitative biomarkers of neonatal IC maturation and white matter injury across both ultra-high-field and conventional clinical 3T MRI.

## Data Availability

All data produced in the present study are available upon reasonable request to the authors.To facilitate the clinical translation and reproducibility of our findings, the source code, trained model weights,
and an executable Docker container are publicly available at https://github.com/SVRTK/7t-brain-analysis.

## Acknowledgments

We thank everyone who was involved in acquisition and analysis of the datasets at the Department of Early Life Imaging at Kings College London and St Thomas’ Hospital. We thank all participants and their families for contributing their time to this research.

This work was supported by a Wellcome Early Career Fellowship (314678/Z/24/Z) awarded to Chiara Casella, an MRC Senior Clinical Fellowship (MR/Y009665/1) awarded to T.A., a Rosetrees Trust grant (2817415) awarded to M.R., a Wellcome Trust Collaboration in Science grant (WT201526/Z/16/Z), an MRC Medium Sized Equipment grant (MC/PC/APP22898), the MRC Centre for Neurodevelopmental Disorders King’s College London [MR/N026063/1], the Wellcome/EPSRC Centre for Medical Engineering at King’s College London (WT 203148/Z/16/Z), the NIHR Clinical Research Facility at Guy’s and St Thomas’, and the National Institute for Health and Care Research Biomedical Research Centre based at Guy’s and St Thomas’ NHS Foundation Trust and King’s College London.

The views expressed are those of the authors and not necessarily those of the NHS, the NIHR or the Department of Health.

## Author contributions

CC and AU contributed equally to this work and share first authorship. CC and AU jointly prepared the manuscript. CC curated and preprocessed the 7T and 3T datasets, implemented and ran the biometry analyses, and performed all other statistical analyses. AU defined the parcellation and analysis protocol, developed the segmentation pipeline, and run segmentation and metric extraction for all datasets. LD contributed to preparation of the training datasets. JWM and BC contributed to curation and processing of the 7T datasets. EG contributed to curation and processing of the 3T datasets and provided support with implementation of the normative modelling. PB, PDC, IT, CDC, and DG contributed to acquisition of the datasets. SA contributed to definition of the parcellation and biometry protocols. MD and JOM contributed to the analysis. SJC provided the 7T datasets. JVH provided the 3T datasets. MAR contributed to definition of the parcellation protocol, the analyses, and study design. SM provided the 7T datasets. TA provided the 7T datasets and supervised the project. All authors reviewed and approved the manuscript.

## Footnotes

2 https://sheffieldml.github.io/GPy/

